# Transmission Dynamics of the COVID-19 Epidemics in England

**DOI:** 10.1101/2020.06.30.20143743

**Authors:** Yang Liu, Julian W Tang, Tommy TY Lam

## Abstract

The ongoing COVID-19 pandemic has caused a tremendous health burden and impact on the world economy. As one of the European countries experiencing one of the worst COVID-19 epidemics, the UK government at the end of March 2020 implemented the biggest lockdown of society during peacetime in British history, aiming to contain the rapid spread of the virus. While the lockdown has been maintained for seven weeks in UK, the effectiveness of the control measures in suppressing the transmission of the disease remains incompletely understood. Here we applied a Bayesian SEIR (susceptible-exposed-infected-removed) epidemiological model to rebuild the local transmission dynamics of the spread of COVID-19 in nine regions of England. We found that the basic reproduction number (*R*_0_) in England is relatively high compared with China. Our estimation of the temporally varying effective reproduction number (*R*_*t*_) suggests that the control measures, especially the forceful lockdown, were effective to reduce the transmissibility and curb the COVID-19 epidemic. Although the overall incidence rate in the UK has declined, our forecasting highlights the possibility of a second wave of the disease in several regions, which may be currently underway in one of the cities there (e.g. Leicester, East Midlands). This study enhances our understanding of the current outbreak and effectiveness of control measures in the UK.

## 1 Introduction

The unexpected emergence and outbreak of the coronavirus disease 2019 (COVID-19), caused by the severe acute respiratory syndrome coronavirus 2 (SARS-CoV-2) (Zhou et al., 2020; Lu et al., 2020a; Lam et al., 2020), has become an ongoing and serious threat to the global health and economy. Early cases of the disease were reported in Wuhan city, China in late December 2019 (Li et al., 2020; Huang et al., 2020a; Wang et al., 2020). Since then, the geographical diffusion of the disease was expedited by the return-to-home migration during the Chinese new year, which led to the reports of a number of successive outbreaks in other provinces of China (Jia et al., 2020; Kang et al., 2020).

Regarding the severe epidemic of COVID-19 in China, the World Health Organization (WHO) declared a Public Health Emergency of International Concern (PHEIC) on 30 January 2020. Although great efforts were taken to contain the disease, with only few imported cases initially reported in Europe and North America in early February 2020, the outbreak soon became global. Outbreaks were reported almost simultaneously in Lombardy, Italy (Grasselli et al., 2020); Daegu, South Korea (Shim et al., 2020) and Qom, Iran (Safavi et al., 2020), which quickly spread to neighboring countries. This resulted in the WHO characterizing COVID-19 as a pandemic on 11 March 2020.

The first confirmed case in the UK was identified in York on 31 January 2020. This was followed by several cases sporadically reported between 1 February and 27 February 2020. Most of these early cases had a clear overseas travel history and they were quarantined and received immediate supportive care. As of 14 February 2020, eight of the nine confirmed cases had recovered. However, the number of confirmed cases in the four nations (England, Scotland, Wales and Northern Ireland) of the UK began to increase rapidly from 28 February 2020.

Initially, London was the most severely affected, where the confirmed number of cases accounted for almost one-third of the total in England by 31 March 2020. Local transmission chains were identified between large cities and neighboring towns and rural areas in almost every region of the UK by the end of May. The death toll also increased along with the number of cases, resulting in the UK overtaking Italy as the country with the highest death toll in Europe and the second highest in the world on 5 May 2020. As of 24 June 2020, there are over 306,000 confirmed COVID-19 cases with almost 43,000 deaths in the UK.

Although the majority of infected patients show only mild symptoms (Chen et al., 2020), including fever and cough (Guan et al., 2020), some patients develop critical symptoms after the hospital admission with likely immune-mediated and aggravated disease (Ye et al., 2020). No effective treatment options had been definitively identified at that time, as demonstrated by well-designed randomized control trials (Zhai et al., 2020), and clinical trials of candidate SARS-CoV-2 vaccines were still in their early stages (Zhu et al., 2020). Thus, the lack of pharmaceutical interventions, together with the high transmissibility of the virus was clearly exacerbating the COVID-19 pandemic in the UK and elsewhere (Huang et al., 2020b).

In addition, it is still unclear whether and how SARS-CoV-2 will circulate and interact with the other seasonal human coronaviruses, in the UK and globally, and to what extent it may become seasonal (HCoV-229E, HCoV-NL63, HCoV-OC43 and HCoV-HKU1) (Liu et al., 2020; Shi et al., 2020; Xie and Zhu, 2020; Yao et al., 2020). Nevertheless, given the ongoing COVID-19 activities in tropical regions, it is now very unlikely that the current UK epidemic will naturally end during summer. Therefore, identifying effective, practical and economic public health interventions both for now and in the future will be critical to rapidly contain the spread of the virus and alleviate the pressure on healthcare systems.

In China, the government banned all transportations to and from Wuhan on 23 January 2020 and subsequently closed the border of remaining cities in Hubei province. Similar measures aimed to reduce human mobility were issued in other Chinese cities and have been shown to effectively mitigate the spread of the infection (Kraemer et al., 2020; Tian et al., 2020). In Europe, Italy first implemented a national lockdown on 11 March 2020. This was followed by Spain on 15 March and France on 17 March, and finally the UK on 23 March (https://www.gov.uk/government/speeches/pm-address-to-the-nation-on-coronavirus-23-march-2020). All these measures eventually proved effective in curtailing local COVID-19 outbreaks in these countries, i.e. by reducing the effective reproduction number below 1 (Gatto et al., 2020; Aleta and Moreno, 2020; Kwok et al., 2020).

Therefore, exploring the transmission dynamics of the SARS-CoV-2 and investigating the effectiveness of various control measures is important to acquire better understanding of this ongoing pandemic to develop and improve public health intervention policies. Although there have been a few studies that mainly focused on China, Continental Europe and North America (e.g., Wu et al., 2020; Zhang et al., 2020; Pan et al., 2020; Kucharski et al., 2020; Liang, 2020; Linka et al., 2020; Leung et al., 2020; Yang et al., 2020), few studies have analyzed the UK COVID-19 epidemic specifically, regarding the local transmission dynamics and evaluation of the control measures.

Here we applied a Bayesian SEIR (susceptible-exposed-infected-removed) epidemiological model that incorporates internal migration data and the regional daily number of laboratory-confirmed cases to reveal the local epidemic progression of the COVID-19 in nine regions of England, including East Midlands, East of England, London, North East, North West, South East, South West, West Midlands, and Yorkshire and the Humber. The regional basic reproduction number (*R*_0_) and temporally varying effective reproduction number (*R*_*t*_) were estimated by a sequential Monte Carlo method to identify the effectiveness of control measures. We also provide forecasts for the number of daily cases for nine regions of England.

## 2 Method

### 2.1 Date sources

Two datasets were used in this study to rebuild the transmission dynamics of the COVID-19 epidemic in England: the daily number of laboratory-confirmed cases between 27 February and 31 May 2020, collected from the publicly available dashboard provided by Public Health England (PHE: https://coronavirus.data.gov.uk/); and the internal migration data collected from the Office of National Statistics of UK (https://www.ons.gov.uk/).

Public Health England (PHE) publishes a case dataset that comprises the number of laboratory-confirmed cases of COVID-19 within different types of administrative areas of England (i.e. region, upper-tier local authority, and lower-tier local authority). The laboratory-confirmed cases were identified in local laboratories of the National Health System (NHS) by testing specimens from people eligible for SARS-CoV-2 testing, according to the national guidance active at that time. The geographical location of each specimen was tracked by the home postcode of the person being tested. If repeat tests were conducted, the date when the first positive test occurred was recorded. Redundant tests from the same person were removed so there was no double record. Cases were aggregated according to the corresponding administrative area. Not all local authorities had complete records, and some administrative regions were too small and did not seem to have significant or continuous outbreaks. Therefore, we focused on regional level data in this study.

In addition, our model only considers local transmissions, therefore cases reported prior to 27 February 2020 (the date when the local transmission was considered to have begun) were excluded. Note that, data was not always up-to-date as some community test results would have been delayed (including care home figures). Therefore, the daily number of laboratory-confirmed cases from all nine regions were collated on 7 June 2020 (one week after the last date of our collected data) and shown in Figure 1. It is likely that the epidemic peak in England was reached on 8 April 2020. COVID-19 cases in London and the North West accounted for the first (18.4%) and second largest (17.6%) number of cumulative cases by 1 June 2020.

**Figure 1:**
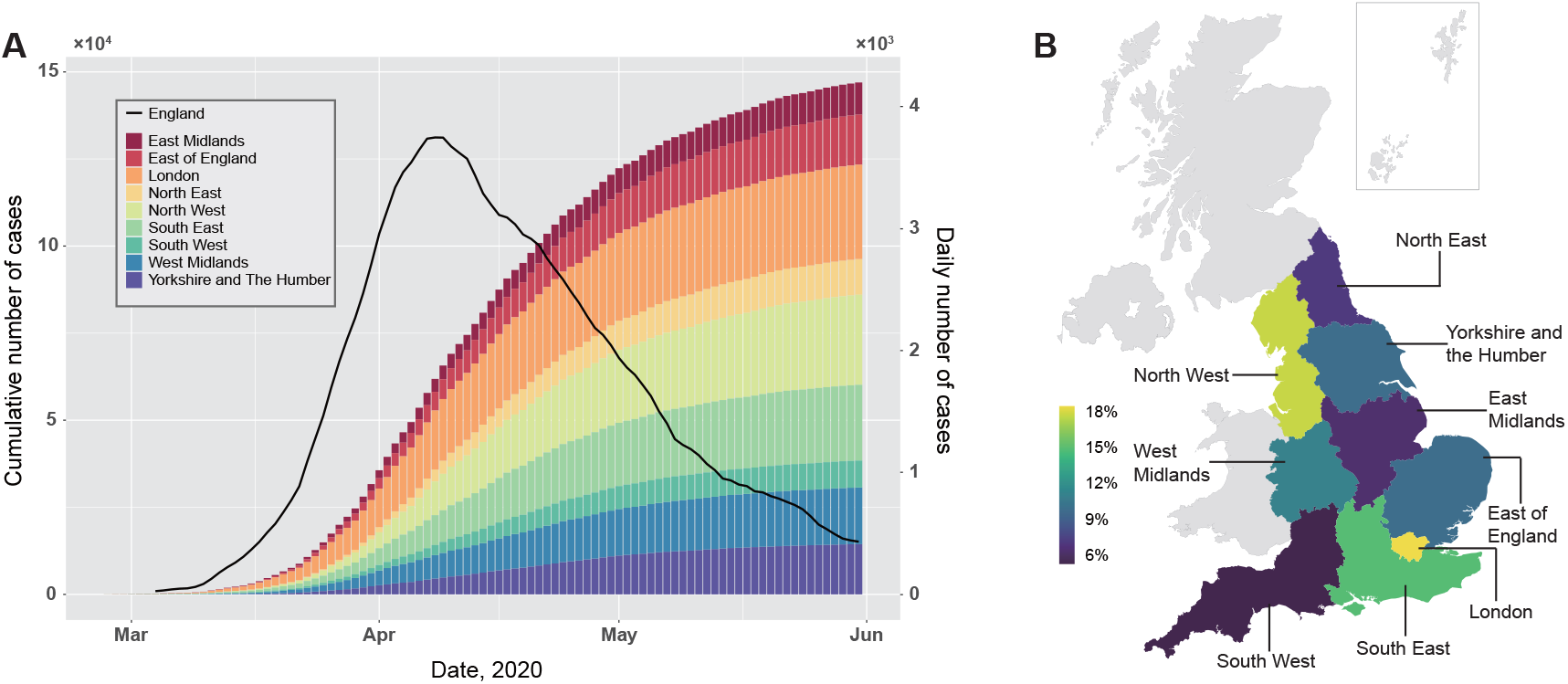
(A). Regional cumulative number of the laboratory confirmed COVID-19 cases in England, and the corresponding national daily number of the confirmed cases, calculated using a 7-day moving average. (B). Geographical distribution of the regional proportion of the cumulative number of the laboratory confirmed COVID-19 cases as of 31 May 2020 in England.

To account for the movement of population between regions in modelling the disease transmission, we used the annual mid-year internal migration data, where internal migration was defined as residential moves across the boundaries of the nine English regions. We used the latest available annual data, from 2018. Inflow and outflow data were aggregated across sex and age. To have a constant population size in the model, the inflow and outflow data involved in the model were transformed so that they were both equal to the mean of the observed inflow and outflow data.

### 2.2 Mathematical model

A SEIR compartmental model, which is widely used in infectious disease modeling to describe the transmission dynamics within a community, was applied here. The model divides the population into susceptible (S), exposed (E; but not infectious), infected (I) and removed (R) compartments and people progress between these disease statuses, which have been clinically described elsewhere (Huang et al., 2020a; Wang et al., 2020; Guan et al., 2020).

Since PHE suggested the citizens with mild respiratory symptoms stay at home and self-isolate, and the COVID-19 patients with non-respiratory symptoms have been reported in some studies (Cai et al., 2020; Lu et al., 2020b), it is likely that there is an unknown proportion of the population in England that have been infected and subsequently recovered without being recognized and tested. Hence, people in the infected (I) compartment are divided into those who are infected and receive test *I*_*a*_ (i.e. those who have severe symptoms and receive medical treatment in a hospital) and people who are infected and do not have a test *I*_*b*_ (i.e. those with mild symptoms and who recovered at home). We know that the proportions of hospitalized and community patients (infected) are significant (Bird et al., 2020; Tang et al., 2020). We assume that a proportion *δ* of the exposed population will enter *I*_*a*_ and a Uniform prior distribution *U* [0, 1] is used to account for the uncertainty about *δ*. The equations of changes in each compartment are set as follows

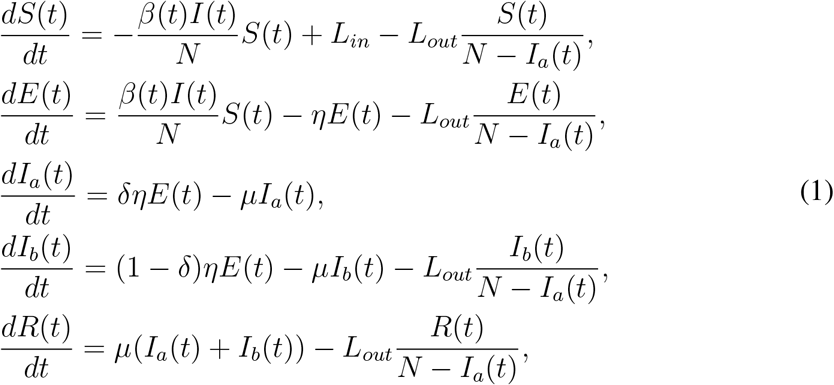

where we assume there is no imported case. In addition to the compartment model, a testing module is added to account for the reporting delay:

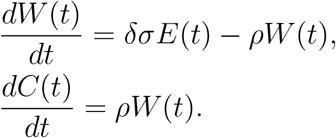

The model is depicted in Figure 2. In this model, *N* is the total population size of the region of interest. *S*(*t*), *E*(*t*), *I*(*t*), *R*(*t*) are the number of susceptible, exposed, infectious, and removed people at time *t. L*_*in*_ and *L*_*out*_ are the inflow and outflow inferred from the internal migration dataset and we assume the inflow and outflow will stop after the national lockdown. We assume that people show symptoms once they enter *I*_*a*_, and *W* (*t*) is the number of people who are waiting for their test result after showing symptoms at time *t. C*(*t*) is the estimated cumulative number of cases. *η* is the rate of being infectious (i.e. the inverse of the incubation period), *µ* is the rate of recovery (i.e. the inverse of the infectious period, which equals the mean serial interval minus the incubation period (Lipsitch et al., 2003)) and 1*/ρ* is the number of days between showing symptoms and receiving test results. We set 1*/η* = 5.2 and 1*/µ* = 2.3 days according to the estimates from a comprehensive study of the early transmission dynamics of COVID-19 (Li et al., 2020) and 1*/ρ* = 4 based on the estimates from Chen et al. (2020). Following Kucharski et al. (2020), we assume a temporally varying transmission rate *β*(*t*) that follows a log-normal sequential update (i.e. log(*β*(*t*)) *~N* (log(*β*(*t−* 1)), *σ*), given a standard deviation *σ*) and the effective reproduction number *R*_*t*_ at time *t* is approximated by (see the Supplementary Method for details)

**Figure 2:**
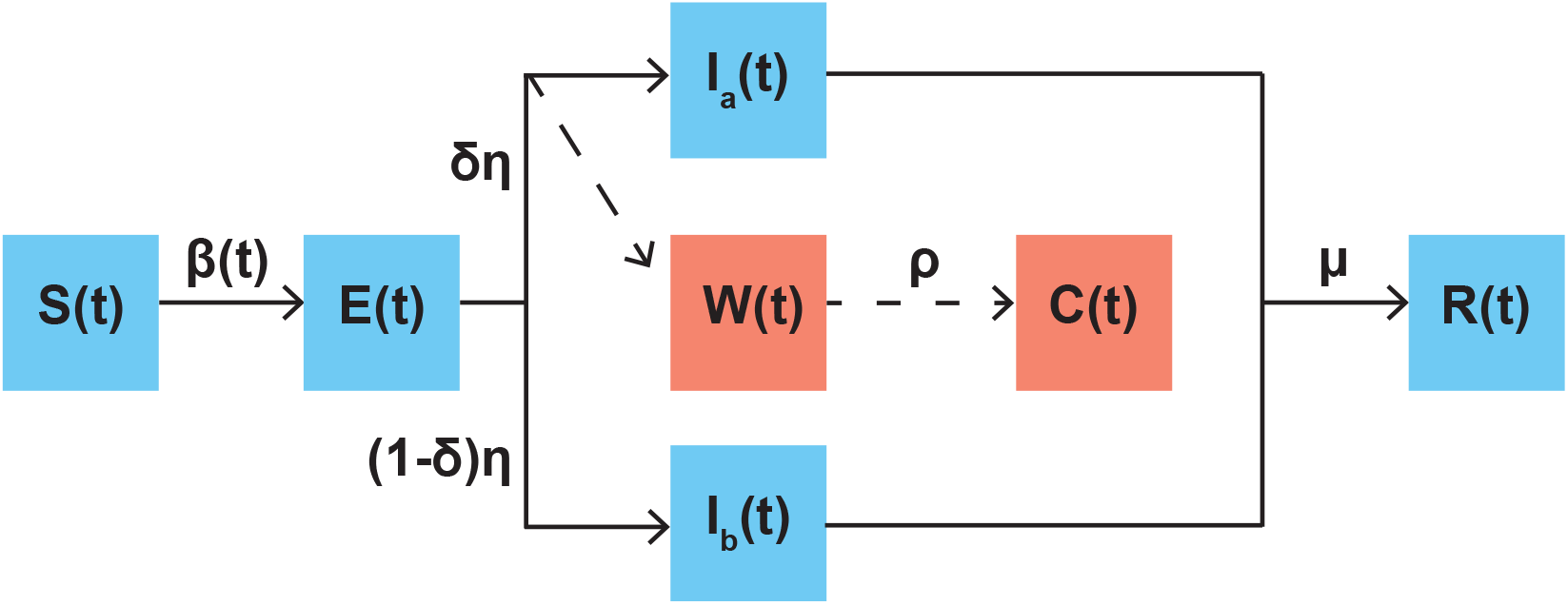
Augmented SEIR structure. The SEIR module and transitions are indicated by blue squares and solid lines; the testing module and transitions are indicated by red squares and dashed lines.

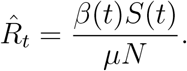

A Poisson generating model is assumed for the daily observed number of the laboratory-confirmed cases *Y*(*t*), given the basic reproduction number *R*_0_ and transmission rate *β*_1:*t*_ from the start to time *t*

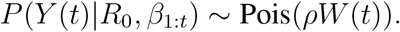

To account for the early period when the virus started to seed in each English region before public awareness, we run a preliminary model that is similar to (1) except with a constant *β* derived from the *R*_0_. The preliminary model assumes that a single infected person enters the region of interest 14 days before the date of the first confirmed local case since 27 February 2020.

The analysis was conducted in R version 4.0. The sequential Monte Carlo method was used to draw samples of *R*_0_ and *β*_1:*t*_ from the posterior distribution, where the optimal standard deviation *σ* that gives the highest likelihood was selected by an exhaustive search.

## 3 Results and discussion

### 3.1 Basic reproduction number

The basic reproduction number *R*_0_ is an important parameter that quantifies the disease transmissibility at the start of epidemic. The estimated basic reproduction numbers *R*_0_ for each of the nine English regions are listed in Table 1 (histograms of the posterior samples are shown in Figure A). All these regions have *R*_0_ between 2.8-3.9, which are significantly higher than 1.

**Table 1:**
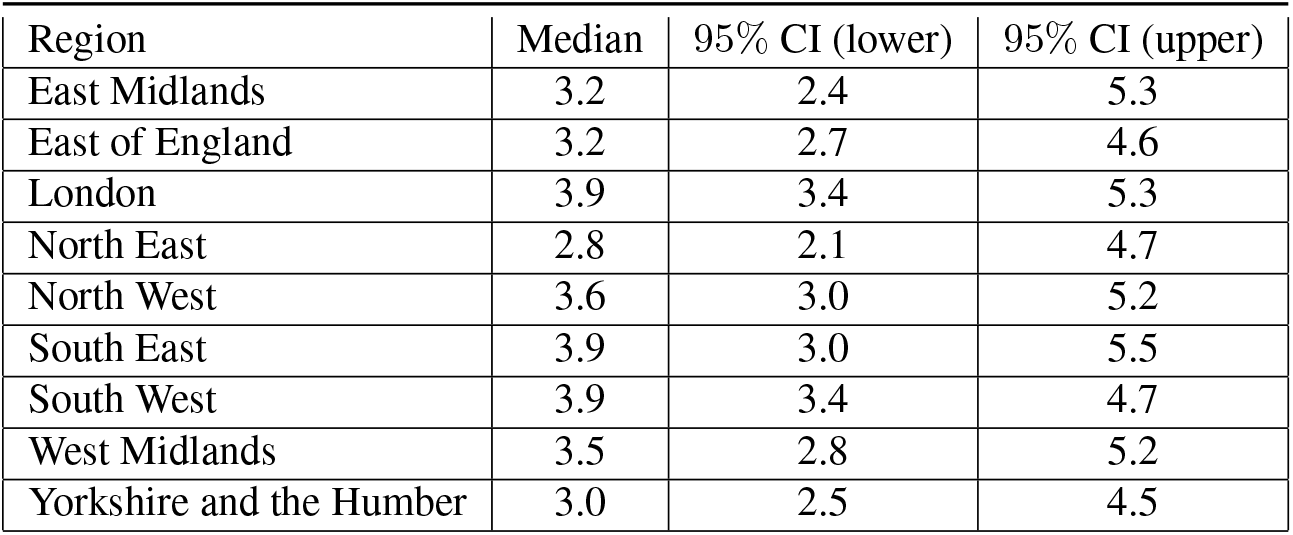
Estimated basic reproduction number *R*_0_ and the corresponding 95% credible interval in each English region.

Notably, we find that the estimated *R*_0_ is positively correlated with the population size in each region. Spearman’s rank correlation is 0.77 which is significantly higher than 0 (*p*-value < 0.05). These estimates are relatively high (seven of nine regions have their medians larger than 3) compared with the early estimate in China by WHO (1.4-2.5), although high *R*_0_ (3.8-8.9) was also reported in China more recently (e.g., Sanche et al., 2020).

Compared with other major airborne viruses, SARS-CoV-2 in England has an estimated *R*_0_ similar to SARS-CoV but significantly higher than MERS-CoV and other human influenza viruses (Chen, 2020). This indicates that the COVID-19 has very strong transmissibility in the early stage and the infected population size will quickly expand without human interventions in all nine regions. Our estimates of *R*_0_ in England are largely consistent with estimates from other major European countries (e.g., Italy: 3.49-3.84; France: 3.1-3.3) (Gatto et al., 2020; Yuan et al., 2020; Distante et al., 2020; Roques et al., 2020). This might explain the rapid spread and high pandemic potential of the COVID-19 when relatively few control measures were implemented in Europe.

### 3.2 Effectiveness of control measures

On 25 February 2020, The UK government announced “Contain-Delay-Research-Mitigate” strategy, but this was only followed by advice that travelers from heavily hit countries should self-isolate. On 12 March, the government started to issue policies for local residents, advising those with respiratory symptoms to self-isolate at home.

After the release of the controversial herd immunity strategy, the UK government advised people against “non-essential” travel and public entertainment venues on 16 March. This was followed by the closure of all pubs, cafes, restaurants, bars, gyms, etc. four days later. On 23 March, a restrictive national lockdown was announced by the UK government and the police force was provided with powers to ensure compliance on 26 March.

Since then, people have not been allowed to leave their homes without limited reasons, gatherings of more than two people are forbidden and social distancing is required in shops. Public transport use has declined significantly during the lockdown period. The control measures began to gradually ease from 10 May when the UK government allowed certain groups of people to work and encouraged outdoor exercise, yet people were warned to “stay alert”. Given these measures, we define the control period as between 12 and 26 March and the lockdown period as between 26 March and 10 May.

The estimated temporally varying effective reproduction number *R*_*t*_ is shown in Figure 3. Almost all nine English regions’ *R*_*t*_ exhibit an overall decreasing trend during the control period. London, West Midlands, and South East show a mostly decreasing *R*_*t*_ during the control period. In the remaining regions, although the *R*_*t*_ increases or oscillates at the beginning of the control period when measures were largely mild suggestions, it started to decrease when the more restrictive and forceful national lockdown was implemented. This reveals the effectiveness of issuing forceful control measures to contain the epidemic. Although, *R*_*t*_ in East Midlands, North East and North West rise after the control period, in all cases it subsequently decreases towards 1 within a week. These transmission dynamics patterns are also consistent with some country-level estimates in literature (e.g., Kwok et al., 2020).

**Figure 3:**
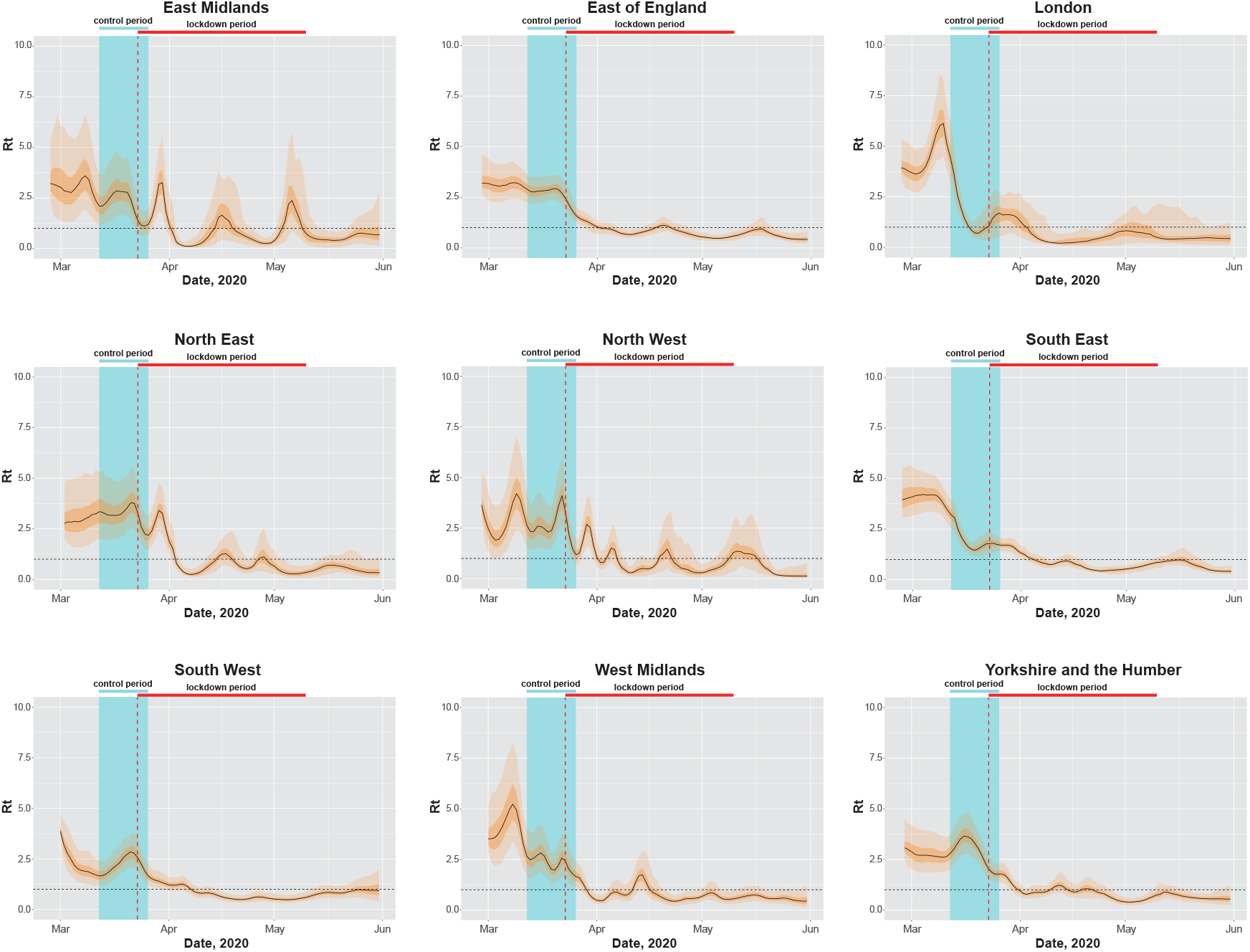
Temporal dynamics of the effective reproduction number *R*_*t*_ estimates based on the daily number of the laboratory confirmed COVID-19 cases between 27 February and 31 May 2020 in nine English regions. The light and deep shaded areas are 95% credible intervals and interquartile ranges respectively. The vertical blue shaded area indicates the control period (12 to 26 March 2020). The vertical red dashed line marks the date of the national lockdown announced on 23 March. The horizontal black dashed line marks the epidemic threshold *R*_*t*_ = 1.

In general, the control measures issued in England during the March were effective to contain the spread of COVID-19 in all nine English regions. This is consistent with a previous study which investigates the effect of the reduction of social contacts on the transmissibility based on surveys (Jarvis et al., 2020). It is noteworthy that the *R*_*t*_ remains slightly under 1 during most of the lockdown period, rather than diminish to 0. This suggests that the local transmissions were maintained, possibly in small towns and communities even though the overall epidemic has been effectively contained, and the virus might circulate between different areas within each English region.

### 3.3 Estimation of the daily number of new cases

To verify the model, we compare the estimated daily number of cases with the observed daily number of confirmed cases. The result is shown in Figure 4 (the estimates of the number of the infected population and susceptible population are shown in Figure B). Although the observed number fluctuates over time, the overall trend of the daily cases is well captured by our model.

**Figure 4:**
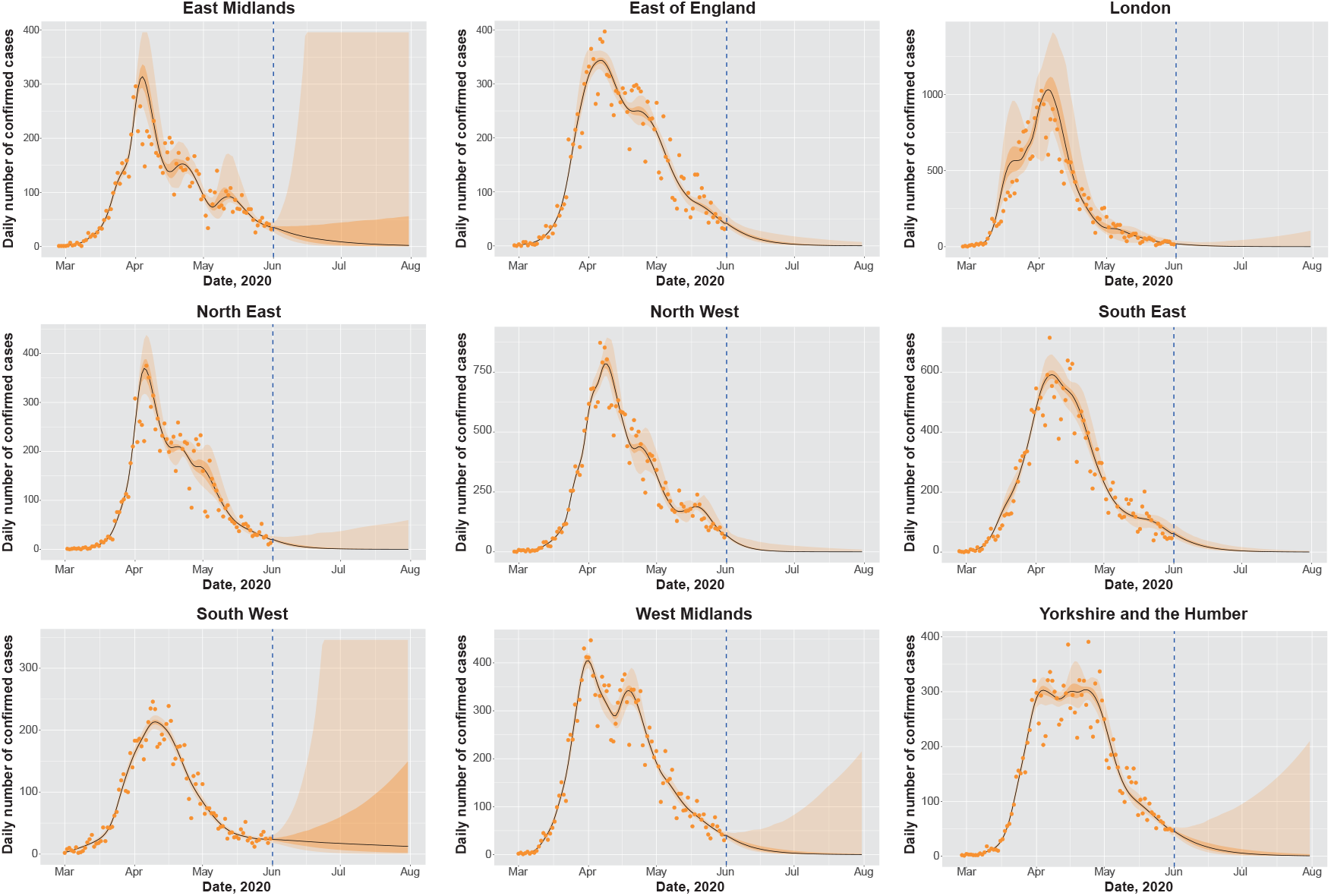
Estimates and forecasting of the daily number of confirmed COVID-19 cases based on data between 27 February and 31 May 2020 in nine English regions. The light and deep shaded areas are 95% credible interval and interquartile range respectively. The dots mark the observed daily number of confirmed cases. The vertical blue dashed line marks the last date of our collected data (31 May 2020). Beyond the dashed line are the forecasting results.

Broadly, the daily number of confirmed cases in the nine English regions achieved their peak in early April. Notably, London is the first region whose daily number of new cases rapidly rose in early March. Our findings highlight the serious epidemic in London during the early phase. This might be due to the fact that London receives the highest proportion of inbound visitors from other English regions and is the most densely populated area among all English regions. In addition, a constantly high daily number of cases was maintained in Yorkshire and the Humber during April, as more infected population may have been seeded prior to lockdown, resulting in the transmission to their household contacts during the lockdown. This may explain the delay in the reduction of *R*_*t*_. The daily number of cases in two regions of the Midlands rose shortly in late April, yet declines are observed from the beginning of May. The daily number of cases in the remaining regions declined during late April and May. These results correspond to our previous estimates that Rt was generally below 1 during the lockdown period. This again indicates the effectiveness of the control measures in all regions.

### 3.4 Forecasting potential second wave outbreaks

In our model forecasting, we assume that *R*_*t*_ stayed constant after 31 May and we use the posterior samples of *R*_*t*_ on that day to forecast the daily number of confirmed cases in June and July. Since the lockdown has been lifted after 01 June, we assume that the inflow and outflow of the population restart. Relevant results are shown in Figure 4.

Although the daily number of new cases is expected to decrease in most regions, it is estimated that, except the East of England, North West and South East, where *R*_*t*_ remains significantly lower than 1, other regions may still witness a second wave of outbreaks.

In particular, the East Midlands, South West, West Midlands, and Yorkshire and the Humber may experience a rebound of incidence after June, as projected by the upper 95% CI of the daily number of new cases (which may go up to 40% and above of the number at the March/April peak).

Notably, Leicester (of East Midlands: https://www.bbc.co.uk/news/uk-england-leicestershire-53105336) and Cleckheaton (of Yorkshire: https://www.bbc.co.uk/news/uk-england-leeds-53105266) recently reported surges of cases, which corroborates our model forecast. Moreover, since the estimated Rt in the South West significantly covers high values on 31 May, it will likely maintain a relatively large median number of infected population until August (more than 100 individuals). While the UK government has been considering lifting some control measures to restore the economy, special attentions should be paid to the regions with the risk of the second wave outbreak.

## 4 Conclusion

Our study demonstrated the use of a Bayesian SEIR model to reconstruct the transmission dynamics of the COVID-19 in nine English regions. Although the true dynamics of the transmission of the COVID-19 is a complex process, the estimated daily number of cases follows closely with the trend of the observed daily number of cases, indicating the validity of our model. Our findings show that the basic reproduction number in England is generally higher than China but in line with some major European countries. The effective reproduction number estimates present a temporally varying trend of the transmissibility. Our results suggest that the transmissibility of the COVID-19 was effectively reduced by the control measures adopted by the UK government. This leads to a decline of the number of infected population during May. Notably, although critics may argue that the restriction of the free movement of people violates basic human rights when the milder measures like social distancing can be equally useful, such strict measures within a national lockdown were efficient to contain the transmission in some regions. The forecasting data highlight the possibility of early secondary outbreaks, so close monitoring of the rate of transmission and *R*_*t*_ are still required even after lockdown measures are lifted.

## Data Availability

The data is publicly available.

## Acknowledgements

The authors thank Robert J.B. Goudie for helpful discussions of this work. Yang Liu was supported by a Cambridge International Scholarship from the Cambridge Commonwealth, European and International Trust. Tommy TY Lam was supported by The National Natural Science Foundation of China Excellent Young Scientists Fund (Hong Kong and Macau) (31922087).

## Ethics approval and consent to participate

Since no individual patient’s data is identifiable in this study, the ethical approval or individual consent is not applicable.

## A Supplementary figure 1

**Figure A:**
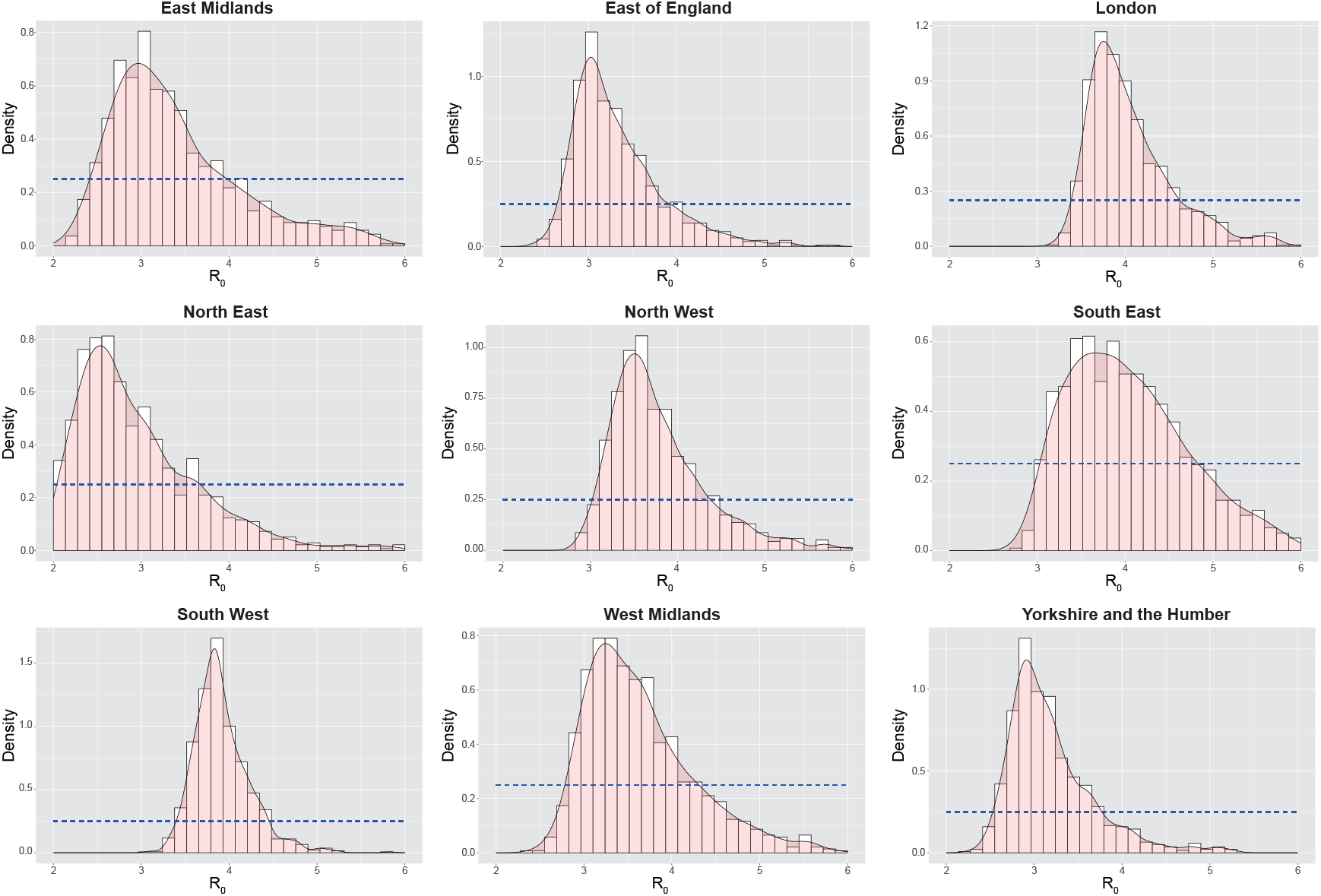
Histogram and the empirical density of the posterior samples of the basic reproduction number *R*_0_ based on the daily number of cases between 27 February and 31 May 2020 in nine English regions. The horizontal dashed line is the prior density.

## B Supplementary figure 2

**Figure B:**
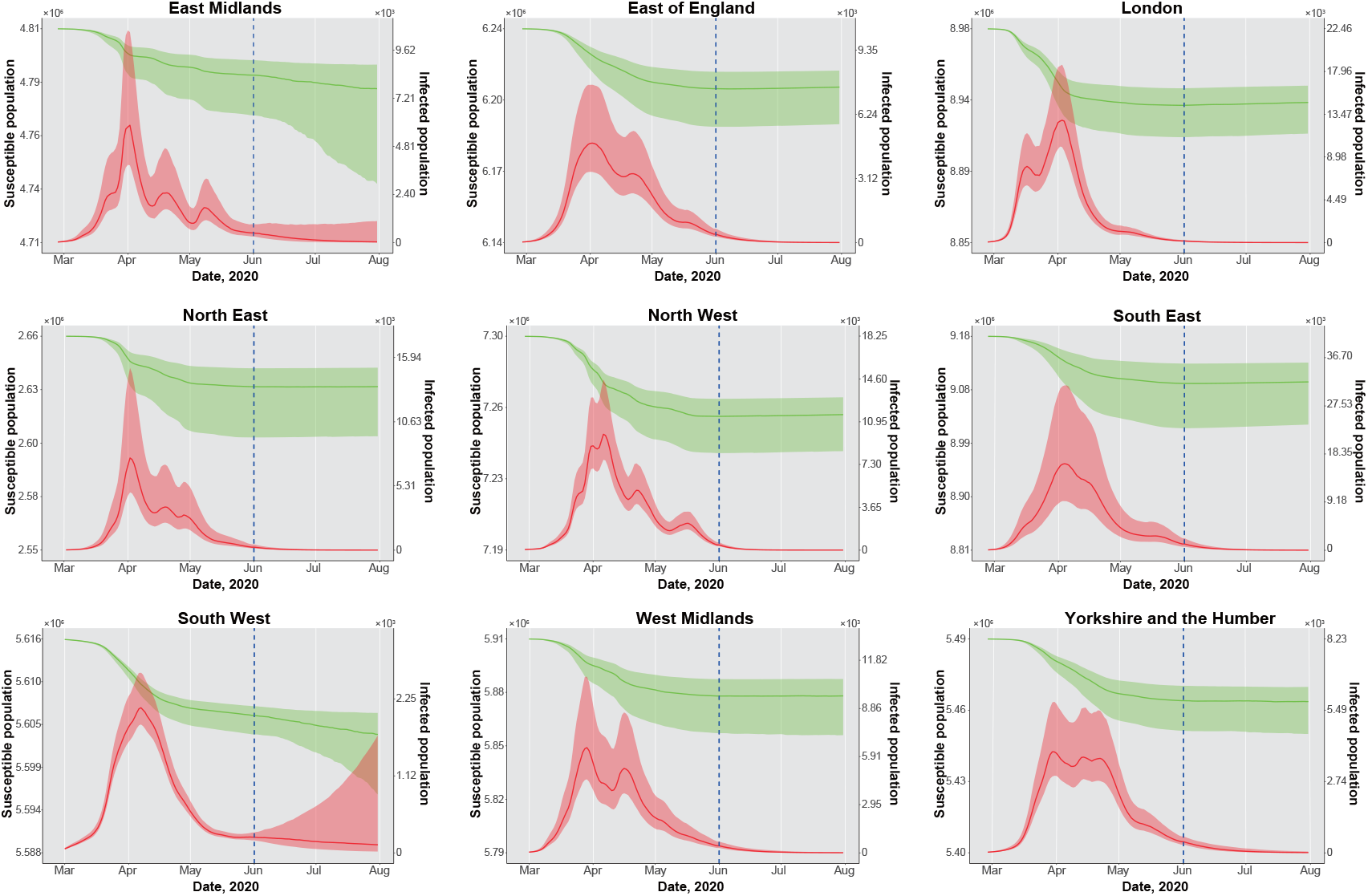
Estimates and forecasting of the number of susceptible population (green) and infected population (red) defined as the sum of individuals in compartment *E* and *I*. The shaded areas are the interquartile range. The vertical blue dashed line marks the last date of our collected data (31 May 2020). Beyond the dashed line are the forecasting results.

## C Supplementary Method

In this supplementary method, we discuss the formula to estimate the effective reproduction number *R*_*t*_ at time *t*. In the main text, we build the SEIR model as:

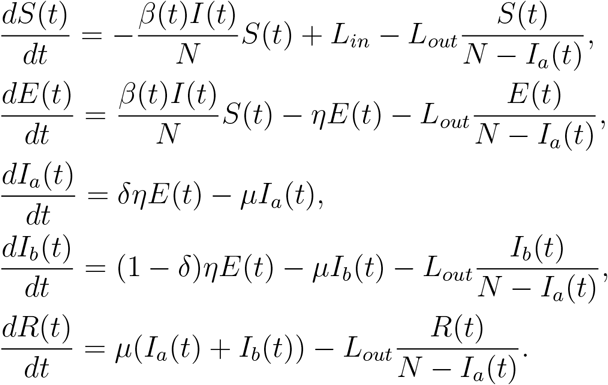

We apply the next generation matrix (van den Driessche and Watmough, 2002) to calculate the *R*_*t*_. Given the infected compartments *E, I*_*a*_, *I*_*b*_, we have

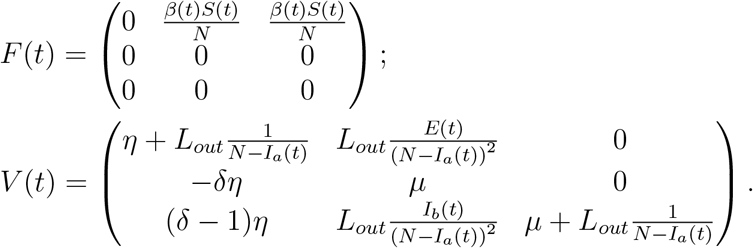

Note that, Figure B shows that the proportion of susceptible population *S*(*t*)*/N* for each region is very close to 1 during the whole period of study. The daily number of inflow and outflow are also far smaller than the total number of the population. To simplify the calculation, we can therefore approximate the matrix *V* (*t*) by

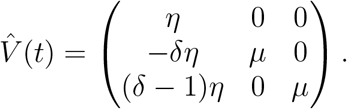

Then we have the approximated next generation matrix:

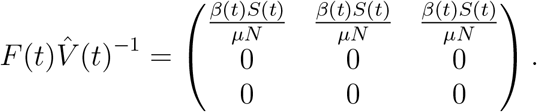

Since the effective reproduction number is the spectral radius of the next generation matrix (i.e., the eigenvalue which has the largest absolute value), then we have

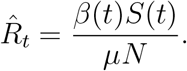

